# Enhancing the Implementation Process of Trachoma Interventions Using Design Thinking Approach in Tanzania: A research protocol to identify the novel strategy in a Trachoma Persistent District

**DOI:** 10.1101/2024.04.16.24305886

**Authors:** Innocent Semali, George Kabona, Yohanna Mshalla, Columba Mbekenga, Deodatus Kakoko, Adelah Sariah, Ambakise Mhiche, Moshi Ntabaye, Godwin Ndosi, Alex Mwijage, Anitha Kemi

## Abstract

**Background:** Trachoma is among the major causes of blindness affecting approximately 8 million people globally. The disease is most prevalent in rural populations with poor access to clean water, health care, and poor environmental hygiene. Current effective efforts to curb the disease include SAFE (surgery, antibiotic treatment, facial hygiene and environmental change) interventions which have shown evidence of real-world effectiveness in the control of trachoma. With the use of SAFE, WHO, governments and other stakeholders vowed to eliminate trachoma by 2020. Though by 2020 people at risk of trachoma had dropped by 90%, few countries including Tanzania were yet to achieve the 2020 goal. Tanzania remained with ten districts highly endemic to trachoma despite regular implementation of the SAFE interventions, suggesting wicked trachoma implementation problems that demand taking different approach to address its complexity. Such an approach will be design thinking to enable identification of effective novel SAFE implementation strategy that is human centered that enables the understanding of those at risk of Trachoma consequently improving access and the utilization of SAFE interventions.

**Aim:** This study aims to gain an understanding of the challenges experienced by communities to access SAFE interventions in the Trachoma endemic areas including stakeholders enabling the understanding of their perceptions, feelings, and behaviors regarding the trachoma implementation interventions. Consequently enabling the designing of effective and innovative human-centered approaches to enhance the implementation of SAFE strategies in the endemic communities.

**Methods:** the study will be implemented in two villages in Manyara region where Trachoma is persistent despite several rounds of mass drug administration (MDA) over several years. It is a design think approach with five phases iteratively. The study will be in two main phases staring with rapid assessment which will employe Trachoma implementation documents and qualitative interviews of selected stakeholders. Its aim will be to understand the ongoing implementation of SAFE interventions performance, promoting and also impeding contexts. Phase two will then follow and will employ five-phased design thinking approach to comprehend the SAFE target population. The design thinking will start with empathy phase, followed with define, ideation, prototyping and finally testing the resulting innovations. As an iterative process each of the subsequent phases will be informed by the previous phase.

The rapid assessment will identify challenges that need exploration, refine research methods and tools, and finalize selection of teams and stakeholders to be involved in the study. The empathy phase will involve obtaining information to gain a deeper understanding of the SAFE target population. Data collection methods during this phase will include qualitative interview, observations, workshops, taking photos and videos. The define phase will involve analyzing information obtained from the empathy phase to develop actionable problem statements that will provide guidance to the next phases. During the ideation phase, in collaboration with the SAFE target population and stakeholders, the research team will formulate possible solutions to address each of the identified problem statements from the define phase. In the prototype phase, the researchers, the SAFE target population and other stakeholders will work together to create an innovative product identified from possible solutions in the ideation phase, which has a high probability of solving SAFE implementation challenges. The final test phase will involve testing the innovative interventions identified in the prototype phase to assess desirability, feasibility, and viability among the SAFE target population follow-up interviews, and observations. A product with such with such qualities of desirability, feasibility, and viability of the innovative product would then passed for future roll out.

Ethical clearance will be obtained from the Hubert Kairuki Memorial University (HKMU) Institutional Research Ethics Committee and permission to conduct the study will be obtained from relevant local authorities. Informed consent will be sought from local authorities and participants before any data collection round. Anonymity and confidentiality will be observed during and after data collection round.

**Author summary:** Trachoma is leading causes of blindness affecting approximately 8 million people globally. The disease is most prevalent in rural populations with poor access to clean water, health care, and poor environmental hygiene. It is an eye infection caused by Chlamydia trachomatis which is transmitted from eye to eye through direct or indirect transfer of eye and nose discharges of infected person to uninfected persons. Most of the infection among children 1-9 years and their mothers, however blindness occurs among those 20 years and above. Efforts to control trachoma were intensified in 1996 when Global Elimination of Trachoma by 2020 (GET2020) was declared through surgery for people with eye trachomatous trichiasis, mass drug administration for those at risk of infection living in endemic districts, face washing and adequate environmental hygiene with acronym SAFE.

Assessments at the end of the time frame in 2020 there was around 92% reduction in the number of people at risk of Trachoma. Thus, about ten percent of the countries had not achieved the goal including Tanzania where ten districts were experiencing Trachoma recrudescence or persistence as global problem despite several rounds of mass administration of Azithromycin. Thus, there was an implementation failure which by using design thinking approach will lead to identification of an innovative strategy that will address this implementation problem. It is funded by Bill and Melinda foundation and ethical clearance will be obtained from the Hubert Kairuki Memorial University in Dar es Salaam Tanzania.

## Implementation gap

Trachoma is contagious disease of the eyes that spread through contact with the eyes, eyelids, and the secretions from the nose or throat of an infected person and accounting for about 8 million cases of global blindness[1]. World Health Organization grades Trachoma into five stages starting with active infectious stage the Trachomatous inflammation also known as follicular (TF). The second stage is Trachomatous inflammation intense (TI) and the third stage is Trachomatous scarring (TS) which is the presence of scarring in tarsal conjunctiva. The fourth stage is Trachomatous trichiasis (TT) and the fifth and last stage is Corneal Opacity (CO) characterized by distorted vision including blindness[2]. Globally, about 84 million people are suffering from active Trachoma, while 7.5 million have trichiasis and 8 million are blind and are in Sub-Saharan Africa.

Blinding trachoma remains a significant public health problem despite concerted efforts to control it dating centuries back. Such efforts include evidence of antibiotic effectiveness in the 1950s that was further corroborated in 1990 when Azithromycin was verified as highly effective against trachoma[3]. Current intervention developments include the promotion of novel interventions whose main constituent included a synergistic strategy that was given the acronym SAFE meaning S (surgery) for tertiary prevention of eye deformities, and A (antibiotic treatment) as secondary prevention, F (facial hygiene) and E (environmental change) which focus on environmental hygiene[4]. SAFE, the integrated strategy was further field-tested in several countries revealing a highly significant effect in the control of trachoma [5, 6]. Global consistency of the real-world effectiveness of SAFE was also positively corroborated by evidence that emerged from China where before the 1980s trachoma prevalence ranged from 50% to 90% but after the mid-1980s which coincided with the rollout of integrated SAFE strategies, trachoma prevalence dropped to less than 10%[7]. In 1996 WHO with partners jointly declared to achieve Global Elimination of Trachoma by 2020 (GET 2020), through the global alliance partnerships and use of Azithromycin in the **SAFE** trachoma control strategy)[8].

Following SAFE real-world effectiveness multiple stakeholders and Governments ratified and embarked on the goal of eliminating trachoma as a public health problem by 2020. Consequently, by 2020, the population at risk of trachoma had declined by 90% while out of 38 countries endemic to trachoma, 27 countries including Tanzania remained endemic despite the concerted SAFE strategies and the expiration of the timeframe [9]. Thus, currently in Tanzania Trachoma remains geographically clustered among some of the difficult to reach populations[10]. Failure to achieve the 2020 goal in Tanzania and some countries, signaled wicked implementation problems with compelling need to comprehend the implementation barriers and contexts of implementing SAFE trachoma control in the remaining countries including Tanzania. This will enable Tanzania adopt more versatile implementation promotion methods to facilitate achieving the next goal defined in the roadmap 2021-2030. This sets 2030 as the new target timeframe for the global elimination of trachoma in those countries[11].

Resetting the 2020 timeframe coincided with the SDG contexts whose one of the critical aim included to increase knowledge through research, and extend partnerships to increase service coverage, specifically on facial cleanliness and environmental improvement to reduce trachoma transmission[12]. Thus some of the implementation hurdles could be solved with innovative research strategies that will successfully address the wicked implementation challenges. In response to these issues, HKMU responded to the Bill and Melinda Gates Foundation’s (BMGF) call for a research proposal addressing implementation challenges affecting NTDs including Trachoma. HKMU has been awarded BMGF grant to enable making a contribution towards addressing the Trachoma implementation challenges prevailing in Tanzania. HKMU will jointly work with the Ministry of Health (MoH) and the Muhimbili University of Health and Allied Sciences (MUHAS) and the University of Dar es Salaam (UDSM) to undertake this awarded research project to create a novel intervention strategy to facilitate overcoming the Trachoma intervention implementation hurdles. Thus enhancing the implementation of Trachoma interventions in an area where there is Trachoma persistence despite the regular control efforts including MDAs.

A recent Lancet editorial on the WHO NTD Roadmap 2012–2020 observed that though 600 million people benefited from existing Trachoma interventions, making further progress will require focusing on the needs and rights of marginalized people in low-income countries[13]. This further asserts that the problem being experienced in the implementation of Trachoma interventions is wicked as it relates to poverty, ignorance, health system, communities, cultures, and politics among others. As a wicked problem, it will require an implementation research and empathic approaches to enable making further progress in the implementation of Trachoma interventions. It will enable understanding communities, their desires, their needs, perceptions, and technical feasibility of SAFE leading to the creation of human-centered innovative intervention strategy to overcome the prevailing Trachoma implementation hurdles. Henceforth, an empathic understanding of the SAFE target populations shall enable successes in addressing the wicked implementation challenges in the process of Trachoma control.

## Problem statement

In 1999, Tanzania successfully piloted mass administration of Azithromycin (MDA) in the SAFE strategy in a few districts[14]. The evidence justified SAFE implementation in 43 trachoma endemic districts identified from the 2002-2004 baseline surveys of 50 districts that had TF prevalence ≥10% according to WHO guidelines for MDA. Subsequent surveys including those from 2012 to 2014 among 31 districts mapped as part of the Global Trachoma Mapping Project (GTMP), MDA was rolled out to 95 districts endemic to trachoma[15]. These were followed by the NTD program impact survey in 2018 which revealed that 63 out of the 71 endemic districts had attained an elimination threshold of TF<5% [16]. Thus, persistent TF and recrudescent TF are major end-game challenges in attaining trachoma elimination by 2030 a timeframe set after 2020.

Following the concerted SAFE strategies, recrudescence and persistence problems were first identified in 2017 and become more established over the 5 years that followed in four districts namely; Longido, Ngorongoro, Kiteto and Simanjiro which had not attained the TF elimination threshold despite multiple cycles of MDA (ranged 8-12 rounds) [17]. In addition, recrudescent TF was observed in 3 districts: Kongwa, Chamwino, and Mpwapwa which also had been implementing MDA regularly since 1999 to date. Sustained recrudescence and or persistence is a reversal of the gains realized by the ongoing investments, further escalating morbidity and disability due to trachoma, limiting school enrollment, poor school outcome and also increased blindness among adults further lowers the productivity while increasing dependence ratio. Consequently, enhancing the poverty cycle and thus compromise the process towards achieving the Sustainable Development Goals by 2030.

It is indeed important to reckon that Trachoma control has reached an endgame challenge characterized by the persistence and or recrudescence consequent to stalled uptake of the standard highly effective SAFE interventions in isolated implementation sites. In response, there has been an observation to undertake in-depth studies to understand in detail the determinants of the observed wicked challenges[9]. The determinants are multi-levels and multi-sectoral including difficult-to-reach communities and are linked with extreme poverty; environmental factors, beliefs and traditions, economic, and health system where many fail to disentangle. Therefore, implementation research will be adopted to study the problem. In this study, we describe a protocol of implementation research in the contexts of design thinking funded by the Bill and Melinda Gates Foundation through Grant Agreement vide Investment reference ID INV-046340. As stated earlier Jointly implemented by Hubert Kairuki Memorial University (HKMU), the Neglected Tropical Diseases Program (NTDs) from the Ministry of Health - Dodoma, Tanzania and Muhimbili University of Health and Allied Sciences (MUHAS).

## Justifications for the implementation problem

The Tanzania Ministry of Health is seeking solutions to the wicked implementation challenges toward achieving the elimination of trachoma as a public health problem by 2030. Most of the remaining districts with persistent or recrudescence TF prevalence above 5% are nomadic populations. Hence the study will be carried out looking for innovative strategies that will facilitate implementation of the Trachoma control interventions to increase the uptake of the control interventions among those vulnerable nomadic population. Thus the study will revisit the implementation strategies among the communities and related contexts in order to gain an empathic understanding of the communities and contexts to create novel interventions that will enable overcoming the wicked implementation problems and consequently achieve effective and efficient strategy for elimination of Trachoma by 2030.

## Study hypothesis

After more than 20 years of SAFE including regular MDA interventions, trachoma is still endemic in 9 districts in Tanzania most of which are clustered within the nomadic dry belt of Arusha, Manyara and Dodoma regions. It is postulated that the limited SAFE intervention implementation success is due to wicked implementation problems emanating from the complexity in intervention design, implementation and contexts(social, cultural, health systems and economic). On that account. such complexity will require using a Design Thinking approach which will enable generating data to facilitate empathic understanding of the nomadic community, and stakeholder’s perceptions, access, thinking and their behaviors towards the Trachoma intervention implementation process. As a consequence, it will enable the acquisition of a human-centered strategy which shall enhance the development of an innovative intervention strategy that will be efficacious to hasten the implementation processes of SAFE among those communities left behind.

## Goal

The goal is to generate empathic data from the target populations and stakeholders that will enable the design and adoption of an innovative strategy to facilitate overcoming the wicked challenges in the implementation process of SAFE trachoma interventions among the nomadic community in Tanzania.

Specific objectives will be aligned to each of the phases of Design Thinking namely *empathy, definition, ideation, prototyping and testing*.

## Specific objectives

1. To conduct rapid assessment and acclimatization to the current SAFE implementation and contexts to the control of Trachoma in the selected study areas.
2. To generate empathic, multidimensional information to enable empathic understanding of the SAFE target populations and stakeholders’ in respect to the implementation of the SAFE Trachoma interventions in the respective area.
3. To conduct empathy mapping to enable the visualization of the empathy data into thoughts, feelings, and actions of the SAFE target population in order give to get a better understanding of the SAFE target population SAFE experiences.
4. To define the core SAFE implementation problems from the empathy data and pitching the problem statement from the perception of the SAFE target population correctly to generate a variety of specific problem statement questions for processing in the next ideation phase.
5. To openly and jointly researchers with SAFE target population and stakeholders jointly in this ideation phase deliberate on the problem statements from the define phase to generate priority solutions that that will then be submitted to prototype phase as potential solutions to the SAFE implementation challenges.
6. To turn the SAFE priority possible solutions from the ideation phase into SAFE solution prototypes or something tangible which can be tested among real SAFE users. It is to identify solutions that have a high likelihood of solving SAFE implementation obstacles as being experienced by the SAFE target population.
7. To test expose the proposed novel SAFE intervention prototype to the SAFE target population for the first time and determine its desirability, feasibility, and viability as a measure that it be effective to address the SAFE implementation problems.
8. To submit proposal to evaluate effectiveness of the novel SAFE implementation intervention strategy among SAFE target population in a wider scale.

## Literature review

Trachoma remains an important public health problem that in the 19^th^ century and much earlier affected all countries. Its prevalence ranged between 50% and 90% and accounted for a significant proportion of preventable blindness[3, 18]. The high prevalence of Trachoma was sustained by the high level of poverty, poor environmental sanitation and poor personal hygiene. It was also fueled and sustained by the interaction effects of people movements, wars and occupation of military troops as well as migration from rural areas to urban and between countries [18]. However, by the mid-1990s, trachoma had been eliminated in Europe and most of North America because of significant developments that led to improved sanitation, water availability and treatment [19].

That notwithstanding, trachoma remained a problem affecting 150 million people mostly children and also among whom 6 million were blind in 48 countries mostly in Africa, Asia and some of South America[20, 21]. The highest prevalence of trachoma was reported in countries South of Sahara including Sudan and Ethiopia and, in some areas, trachoma was hyperendemic as the prevalence among children 1-9 years was more than 50% while about a fifth of the adults was found to have high scarring trachoma[20]. To address the remaining burden of trachoma, in 1997 the World Health Organization announced a strategy to eliminate trachoma as a cause of blindness by the year 2020.

## Causal agent and natural history of Trachoma

Chlamydia trachomatis is the causal agent of Trachoma disease, a leading cause of blindness currently estimated at 8 million cases globally[22, 23]. Chlamydia trachomatis, is an obligate intracellular bacterium isolated for the first time in 1958 despite the existence of the disease for centuries[18]. It is both directly and indirectly transmitted from one person to the other through eye to eye during close contact and also spread by fingers. An indirect spread involves sharing of fomites, clothes, or even playing tools and particular species of flies which also play significant role in the transmission [24, 25]. The disease also commonly occurs in clusters of age, gender, villages, hamlets, households, or even bedrooms and residential areas[26, 27] Trachoma natural history occurs in two distinct phases, phase one or “active phase” occurs largely among children 1-9 years followed by phase two that occurs two to three decades later among adults also known as cicatricial (scarring structural damage). Initially, the early phase is commonly found in children manifesting as recurrent chronic follicular conjunctivitis[28–30]. As recurrent chronic follicular conjunctivitis persists and becomes more severe it leads to conjunctival scarring which results in shortening and distortion of the eyelid. Consequently, the scarring of the eyelids and shortening causes inward-turning, an effect referred to as entropion[31]. The in-turned eyelids causes the eyelashes to come into contact with the eyeball resulting in trichiasis. At this stage, the eyelashes scratch the cornea resulting in scarring that leads to corneal opacification later manifesting as blindness or diminished vision which is abated by surgical intervention.

## Epidemiology and risk factors for Trachoma

Recent estimates in 2022 reported that populations in areas that warranted intense SAFE interventions had declined by 90% while those with Trachomatous trichiasis had declined by 80% revealing very significant gains consequent to the global, national and stakeholders efforts toward the control of Trachoma[32]. This reflects an estimate of 125 million people are at risk of trachoma blindness while 1.9 million are estimated to be blind or have visual impairment due to Trachoma[33]. These current global figures reflect the gains from various global and national efforts to eradicate the disease when compared to 1995 estimates revealed that 150 million people were affected, in 48 countries, and 6 million were blind[3, 20].

As stated above, the infection and prevalence of active Trachoma disease starts early in childhood peaking among children less than five years, declining gradually to a very low percentage among those 21 to 40 years[30, 34]. On the other hand, the prevalence of scarring trachoma (trichiasis) increases gradually from 1.0% among children under five years to 38% among those 40 years and older in endemic areas. It is therefore agreed that young children constitute a reservoir for trachoma with direct effects on those in close proximity especially mothers and other children. Hence this could explain the high risk of trachoma estimated at 3.9 more likely among women compared to males [28]. The phenomenon is further supported by the results of a systematic review which observed that about three-quarter of corneal blindness and trichiasis occurs among women compared to men [35]. Hence, women constitute a high-risk group perpetuated by their responsibilities as caregivers among families calling for concerted efforts to enhance access to SAFE strategies among women thus offsetting this gender inequality. In addition, the likelihood of trachoma infection increases with the presence of nasal and ocular discharges among children and having more than two children in a household [36].

Other risk factors for trachoma include environmental factors supported by the observation that that living in a household that consumed 60 liters of water per day significantly lowered the likelihood of being infected with trachoma[37]. Thus, easy access to water would contribute to frequent washing which would interrupt the transmission dynamics of the causative agent from those infected to those un-infected. This was also corroborated by the findings that those who used less than two liters of water for face washing per day had an increased risk of trachoma infection. The role of water scarcity and endemicity of trachoma is also corroborated by the observation that the risk of trachoma diseases has been reduced significantly as water sources increased [7].

Furthermore, a significant increase in the risk of trachoma was observed where environmental hygiene was poor which included the presence of liquid waste around the house and also the presence of fecal matter in the household proximity [3]. In addition, the risk was significantly elevated where personal hygiene was poor due to lack of face and hands washing in addition to the presence of flies around children’s faces [36]. Also, poor environmental hygiene, the location of the toilet within the family setup was associated significantly with an increased risk of trachoma if it was less than 10 meters away from the living house.

## Interventions to control, Trachoma

While households should always have some optimal amount of water to ensure that life goes on, it will indeed be wise with the same water per capita use economies of scale approach as an interim measure to enhance facial cleanliness in households and especially children and women. That notwithstanding, interventions to control trachoma are complex and should include focusing on the environment, personal hygiene as well as the trachoma causative agent. Following the 1996 Global Scientific meeting to control Trachoma, the World Health Organization WHO Alliance for the Global Elimination of Trachoma by 2020 also referred to as WHO GET2020[8] was initiated. As a prerequisite to achieving this goal, WHO adopted the SAFE strategy to enhance the trachoma transmission dynamic[38] Systematic application of SAFE strategy in China and some other countries recorded a significant prevalence decline among children 1 to 9 years (children_1-9_) from 55% in early 1980 to less than 5% in 2014, in addition trichiasis declined to less than 1 percent [7]. The decline provided evidence that the SAFE strategy was effective and highly capable to achieve the WHO’s ultimate goal of eliminating trachoma by 2020.[38]

## Monitoring progress in Trachoma control

Also indisputable evidence exists supporting the multi-country rollout of the SAFE strategy among the 36 countries that were endemic to trachoma in the late 1990s and early 2000s [9]. The success of the strategy was monitored using the WHO’s ultimate intervention goals and indicators toward achieving the elimination of trachoma as a public health problem [39]. The following are the indicators that include mass drug administrational (MDA) frequency based on prevalence data from community-based on regular trachoma surveillance among children between the ages of 1–9 years (Children_1-9_):

1. SAFE implementation in a country or a geographical area must reduce the number of people with trichiasis through surgery to less than 0.2 percent among people aged 15 years and above in a district.
2. Also, each country must reduce the number of cases of active trachoma (TF) in children of ages 1 to 9 years (children_1-9_) to less than 5 percent of the population of children in any district using Azithromycin to be declared that a country has eliminated trachoma as a public health problem. Furthermore the following is the frequency and duration of azithromycin mass distribution based on the prevalence of clinical signs of trachoma among children ages 1–9 years as determined by population-based surveillance prevalence surveys [40]:

a. If TF among children 1–9 years old is less than 5.0%, MDA is not required
b. If TF among children 1–9 years old is between 5.0% and 9.9%, one year of MDA is recommended, followed by an impact survey at least 6 months following the last MDA
c. If TF among children 1–9 years old is between 10% and 29.9%, three years of annual MDA is recommended, followed by an impact survey at least 6 months following the last MDA
d. If TF among children 1–9 years old is 30% to 49.9%, five years of annual MDA is recommended, followed by an impact survey at least 6 months following the last MDA
e. If TF among children 1–9 years old is above 50%, seven years of annual MDA is recommended, followed by an impact survey at least 6 months following the last MDA. Once TF among children 1–9 years old drops below 5% in a district, the program should wait for two years before conducting a population-based surveillance survey.
3. Facial Cleanliness and Environmental Improvement UIG Hygiene promotion and environmental improvement should be conducted in a community so that, at any given time, 80 percent of the children in the community will have clean faces.

## Outcomes in the control of Trachoma

Whereas38 countries embarked on the rollout of SAFE including mass administration of Azithromycin, only Morocco, Ghana, and Togo were certified by the WHO to have attained the trachoma prevalence elimination threshold as a public health problem [9]. After attaining the respective prevalence elimination threshold, the countries embarked on close surveillance to detect recrudescence of the infection and take appropriate action to control chlamydial re-infection which had also been observed to occur after successful mass drug administration in other countries[9]. However, despite the availability of resources and facilitating contexts, other countries including Tanzania are experiencing both recrudescence and persistence due to wicked implementation problems and hence are indentured to discern the prevailing implementation challenges and contexts timely to make the desired progress.

However suggestion to tame the SAFE implementation wicked problems could be through the acquisition of strategies that are based on the SAFE client’s needs and their desirables and their participation to inform the development of a novel strategy that overcome the SAFE implementation problems thus enhancing access to the interventions [41]. Thus, it is envisaged that successful implementation of SAFE should not be based researchers preconceived notions of what are the needs and solutions but require systemic solutions that are grounded in the SAFE community’s needs, as well as their desirables. Hence such an approach excels because it unleashes innovative solutions based on perceived and felt needs, desires, perceived solutions which is a systemic deployment of design thinking[41].

This study aims to employ a design thinking approach in the context of implementation research to design context specific innovative strategies that will enable addressing the implementation problems stalling the implementation of SAFE interventions in the control of Trachoma among nomadic populations in Tanzania thus, achieving the Trachoma prevalence elimination threshold.

## Methodology

Implementation research is a scientific undertaking towards solving a problem arising from the implementation process aiming at achieving the desired goal. It is now becoming famous in health and is used to address problems arising from the implementation of clinical, public health, or health system interventions[42]. In that case, the implementation research approach will be proper to disentangle complex factors affecting the implementations and also introduce modified or novel interventions or solutions into the program or the health system.

Fundamental to implementation research is to generate an adequate understanding of what, why, and how interventions work in real-world contexts, for this case the Trachoma interventions[43]. With such adequate understanding in the real-world setting, we will find and test in this study the effect of novel strategies to improve the performance of the SAFE interventions. Working in a real-world context exactly implies working with communities that are facing SAFE implementation obstacles resulting to Trachoma recrudescence or persistence. Nevertheless, this study will be working with the communities experiencing Trachoma persistence after optimal administration of SAFE and MDA (mass drug administration) as proven effective Trachoma interventions among nomadic population in Manyara region in Tanzania.

Additionally, real-world contexts envisage working with the population while focusing on an in-depth understanding of the multilevel contexts which among others includes interaction with both primary and secondary stakeholders, social-cultural, legal, economic, political, and the environment. Others will involve public structures including the health systems, government departments, civil organizations as well as other non-governmental organizations. Furthermore, it emerges that implementation research is focused on optimizing community inputs and involvement to ensure the relevance and effectiveness of the interventions [38]. Such an approach is also identified as community-based participatory research with a strategy to ensure researchers study the correct problems using appropriate methods and problem-relevant interventions or advice suggesting alignment with the appropriate community needs.

Implementation research is human-centered and multilevel focusing on people, communities, and health systems supposed to benefit from the SAFE Trachoma interventions[44]. However, to ensure that the communities participates in developing the solutions and adopts the results of the implementation research design thinking approach will be adopted [36]. Design thinking is an approach which is a problem-solving framework with a set of methods starting with empathic understanding of those affected, generation of creative ideas and ultimately testing those ideas and wide-scale adoption [41].

## Study setting and population

The setting of the study will be a village in Trachoma endemic district experiencing trachoma persistence after an optimal rounds of MDA in Manyara region which will be selected by the National Trachoma Control Program in the Ministry of Health in Tanzania. The study population will be all people in the two selected villages whose population are the SAFE target population since they are all at risk of trachoma infection because of the persistence. Other study population will include, NTD program staff at all levels, Council Health Management Teams, Health Facility leaders, and community leaders as primary stakeholders. The research team will be composed of members with different expertise including ethnographers, social scientist, public health experts, epidemiologists, community development experts and also the national and district Trachoma and NTD coordinators. The study will be implemented in phases iteratively, however, at each phase, its inputs and outputs will be the product of collaborative efforts of the multi-disciplinary team, those affected or at risk of trachoma but also they are targeted for mass drug administration (MDA) whom also will be referred to as the SAFE target population. and a team of multi-experts.

## The Approach

Design thinking is a non-linear, open-minded iterative process that our research team will use to understand the SAFE target population, challenge assumptions, redefine problems, and end up with a cutting-edge innovative solution to the SAFE implementation obstacles. Design thinking, brings together what is desirable from a human point of view with what is technologically feasible and economically viable. The pathway to the successful innovative solution is a five-phase journey starting with empathy, then definition followed by ideation, prototype, and finally testing. Finally, the possible intervention will be assessed by the extent of positively it meet needs and desires of SAFE target, its feasibility and its viability. The methodology will iteratively roll over in five-phase, each successive phase building on the preceding phase as follows starting with phase 0:

## Phase 0: Acclimatization and rapid assessment

This phase will include forging strong collaborations with stakeholders especially the Trachoma Program at the Ministry of Health, expertise from the Muhimbili University of Health and Allied Sciences (MUHAS), University of Dar es Salaam(UDSM) and the regional teams. It will also involve document reviews, rapid assessments and consultations as follows:

a. At onset the three universities and the National Trachoma Control Program of the Ministry of Health will meet discuss and agree on implementation modalities as well as the implementation time-frame and other administration needs. The discussions will also identify expertise and experts who will comprise the field research teams. Participants from MUHAS and UDSM will also have regular meetings and capacity building workshops for the researchers which will focus on implementation research and design thinking for at least one week.
b. **R**esearch team will undertake a rapid assessment to generate a clear understanding of the current magnitude of Trachoma and uptake of the interventions. The phase will also facilitate the identification and clear detailed description of the research problem, the implementation scope and limitations, problem analysis and assessment of already available information, and verification of the villages that will be studied. Data sources for this phase will include reviews of secondary data, grey literature, observations, and a limited collection of qualitative data among stakeholders whose theme will be identified in the process.
c. This phase will be culminated with synthesis workshops to enable stakeholders to discuss and deliberate on the results of the rapid assessment. Outcomes will be a common understanding of the challenge to be explored and of the project’s scope and further refinements of the methodology, data needs, implementation contexts and tools for the empathy phase.
d. This study will utilize design thinking to address this wicked SAFE intervention implementation problems. However, there will also be orienting of all the participating researchers on design thinking to make sure they have uniform understanding and hence common practice. During this phase there will be one week training to enable research team acquire such competencies.
e. Implementation in two selected villages will include formation of two field research teams composed of different experts and two research assistants’ in each team. It will also prepare relevant data collection tools that are conducive to the research contexts. The initial preparation of tools will depend on the findings from the rapid assessment. Before the beginning of each subsequent phase there will be a workshop to plan the phase, prepare tools and accessing needs as will deem necessary in that particular phase and the phases will be in the following order:

## Phase 1: Empathy Phase

Empathy will be putting ourselves in the shoes of the communities experiencing SAFE implementation obstacles (also to be referred to as the SAFE target population) and connecting with how they are feeling about their problem, circumstance, or situation. In this case, our team will put itself in the shoes of the SAFE target population. It will also enable us to connect with how they feel about the problem and the circumstances while putting aside our preconceived knowledge, and own assumptions about the world to gain real insight into the SAFE target population and their needs. Essentially, the empathy phase will predominantly employ ethnography qualitative design [45] that will allow prolonged engagement with the SAFE target population within their communities. Two ethnographers’ researchers will lead the research team to undertake the empathic data collection in the respective two study villages focusing on the SAFE target population i.e. what they need, what they want, and how they behave, feel, and think.

### Objective

Undertaking this phase will enable the research team and stakeholders to generate human-centered rich and multidimensional information to understand the SAFE target population and communities. It will enable seeing the real world through the SAFE target population’s eyes, and stepping in their shoes to feel what they might be feeling about their problem, circumstance, desires and perceive the solutions from their intuitions.

The researchers and data collection team will be primed in advance in the process not to be presumptuous or hasty with SAFE target population perceptions and always operate an ignorant mindset to enable painting a clear picture of the SAFE target populations. Therefore, the two ethnographers in collaboration with the research team and other relevant stakeholders will ensure collection of empathic data relevant to SAFE implementation problems and related contexts. It will focus on the generation of real world adequate data that will include observing SAFE target population and their behaviours in the context of their lives and Trachoma control activities. It will also include interactions and interviews with the SAFE target population using variety of data collection methods. Finally it will involve stepping in the SAFE target populations’ shoes in order to eexperience what they experience over the course of implementing the SAFE interventions and the MDA rounds. Various qualitative methods will be used to collect ethnographic data that precisely answers the above questions as follows:

1. **Observing SAFE target population.** The research team guided by the ethnographers will also observe the SAFE target population behaviors and interactions related to SAFE in their environments and other contexts. This will also take place in the villages, schools, places of gathering, homes as well as health facilities. The observations will enable research team understand how SAFE target population utilize or practice the SAFE interventions. In addition, it will reveal their needs, pain points, and even unintended uses SAFE interventions.
2. **Conducting photo (Photovoice)- and/or video-**based studies in SAFE target population natural environments including their interaction sessions with the research team and reflective notes in relation to each photo/video undertaken.
3. **Interviewing SAFE target population** – The ethnographers and research team will purposively select individuals from the SAFE target population and then ask for their insights in an intimate setting where they can respond earnestly to open-ended questions. The interviews will be a combination of individual in-depth interviews (IDIs), key informant interviews (KIIs) and focus group discussions (FGDs) depending on the focus and information to be explored. Kiswahili and local language of the SAFE population will be used during the interviews. A translator will be identified to translate interviews for participants who do not speak or understand Kiswahili language.
4. **Engaging with the extreme SAFE target population.** – Extreme SAFE target populations will be identified to determine the greatest degrees of SAFE target population needs, problems, problem-solving methods, and persons to establish accurate portraits/profiles of users who will interact with our innovative product. The engagement will comprise of FGDs and/or community workshops with various relevant stakeholders.

### 1.2 Empathy mapping

This phase follows and compliments the empathy data collection phase and includes defining the problem and problem statement which must be done in a human-centered manner. This phase will help our research team frame and explain the SAFE implementation obstacles that were identified during the empathy phase so that novel solutions for those problems can be created later by the community stakeholders.

**Objective:** Empathy mapping will enable the visualization of the empathy data into thoughts, feelings, and actions of the SAFE target population in order give a better understanding of the SAFE target population SAFE experiences and can be used to identify potential areas of improvement by capturing their thoughts, emotions, actions and behaviors through a collaborative visualization of with the SAFE target population.

The research team will use the data acquired from the empathy phase to analyze and synthesize it while also assigning it to the relevant quadrant (figure 1) to enable empathy mapping. Eventually, the research team will create an empathy map that will enable one to understand SAFE target population needs by type in the course of developing a deeper understanding of the SAFE target population’s needs. The empathy map (see Box 1) will have four major areas in which the research Team will jointly align the empathy results to respective quadrants guided by the following questions:

1. What did the SAFE target population say? Data collection will focus on writing down significant quotes and important words said by the respondent.
2. What did the SAFE target population do? Data collection will focus on the description of which actions, and cultural norms, practices and behaviors one noticed, inserting pictures or drawings will add value.
3. What did the SAFE target population think? Also, data collection will have to dig deeper to answer some of the arising questions.
4. How did the SAFE target population feel? What emotions might the SAFE target population be feeling? Data collection should record subtle revelations like body language and their choice of words and tone of voice.

The researchers will identify needs based on contradictions between two traits, such as a disconnection between what a user say and what the SAFE target population does. Results from this phase will be passed to the next phase which is **ideation**. Design thinking is an iterative process; it might also require further empathy inputs to make clarifications or complement existing information.

#### Box 1

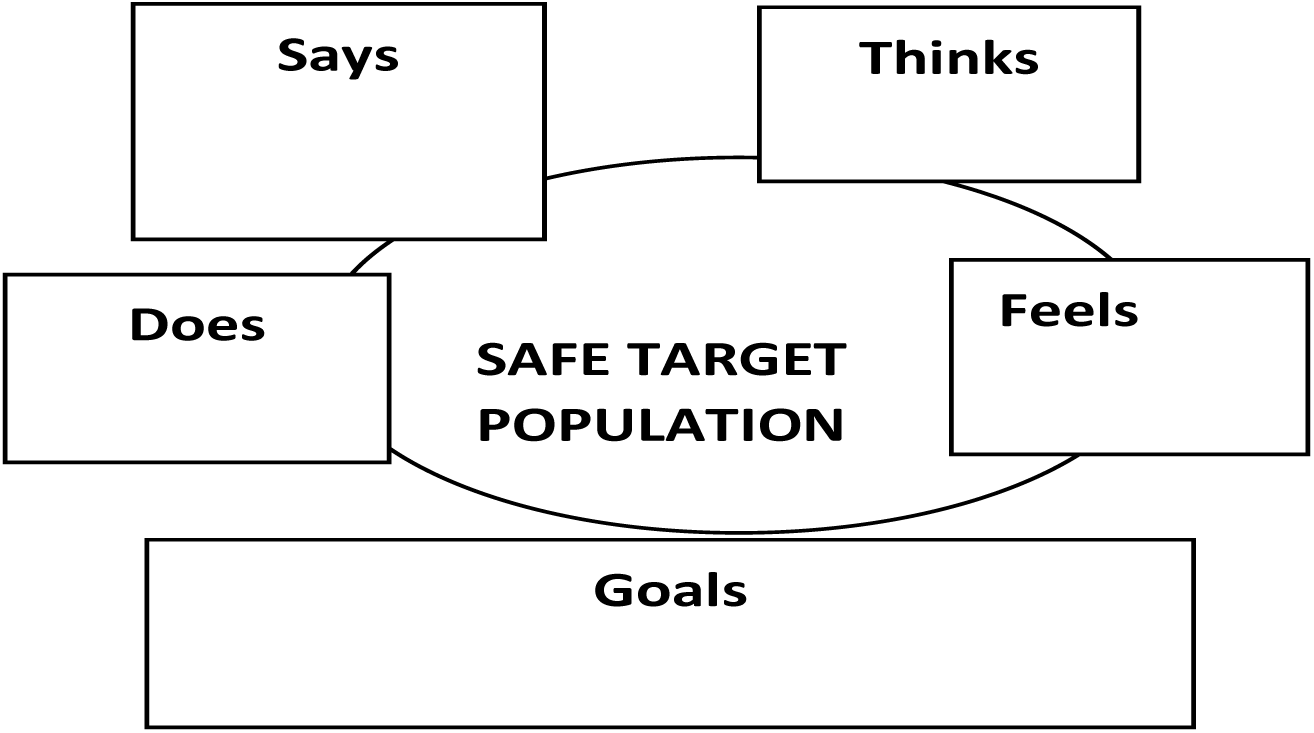
Empathy map.

## Phase 2: Define phase

The define phase will follow the empathy phase where the researchers will take on board all information gathered during the empathy phase, analyze, and synthesize it. In addition, the researchers will shape the information into a problem statement which will act as the main guide towards articulating the problem and providing a clear-cut objective to work towards.

### Objective

To provide meaningful, actionable SAFE implementation statements which will steer the research team in the right direction, helping the research team inaugurate efficiently the ideation process to inform the teams and others what can be expected to be achieved as a SAFE implementation innovative solution.

The define phase is immediately preceded by the empathy phase where the researchers will have learnt lot about the SAFE target population and most importantly their wants, needs, and pain points. It will include understanding who is experiencing Trachoma and SAFE implementation problems, what are the SAFE implementation and Trachoma problems, what is the periodicity and what are the benefits of solving the problem.

Having achieved this, the research team will turn the information from empathy into an actionable problem statement. A problem statement identifies the gap between the current state (i.e. the problem) and the desired state (i.e. the goal) of SAFE interventions among the SAFE target population. The researchers will cast the problem as an unmet need, therefore a good problem statement will be human-centered in this case the SAFE target population and also focus on their needs. Our researchers will frame the problems in actionable format keeping a focus on the SAFE target populations at all times.

All problem statements will always be formatted and focused on the SAFE target population perspective. We will avoid statements that start with “we need to…” or “the SAFE target population should”, we will instead concentrate on the SAFE target population’s perspective: “SAFE target population need…”, and should not be too broad

An example of casting a statement is.

> *“I am a young unemployed father of six children trying to get food and living for my family from a distant village, but I’m struggling because I work long hours and don’t always have time to attend SAFE campaigns and access the interventions. This makes me feel frustrated and bad about myself and my family.”*

Our research team will then condense all the complex data obtained from the SAFE target population into simple, actionable statements. The research team will use among others a method that will help to condense the information. This study will make use of a way to dig deeper into the problem until a pain point is reached. A workshop of the research team, SAFE target population, and co-opted stakeholders will be held which will condense the data. At this stage, empathy data will have been summarized into meaningful, actionable problem statements. These will be actionable statements that have clues to solutions that will be ready to be moved to the next ideation phase.

## Phase 3: Ideation phase

This phase will follow the define phase and will involve generating a broad set of ideas on a given topic arising from the define phase, in a brainstorming workshop of the research team with no attempt to judge or evaluate them. It will in this case represent an important transitional step from learning about the SAFE target population and the SAFE implementation obstacle, as well as coming up with solutions.

**Objective:** To enable research team, SAFE target population, and other stakeholders in a facilitated and judgment-free environment openly generate as many ideas and solutions as possible while addressing each of the selected problem statements from the define phase.

In this phase our research team will rephrase the problem statements into a list of questions of “How might we..” Thus, the team will look at each insight statement that the team created in the define phase and rephrase it as questions by adding “How might we” at the beginning.

After creating the list of “How might we….” We will generate possible answers which will become a catalyst for the brainstorming workshop. Our research team will compile a list of possible answers and subject each to an ideation technique. The following ideation approaches will be employed to generate possible solutions concerning the stated problem: The team (Research team, SAFE target population, and Co-opted stakeholders) decide on which minimum combination of these techniques will be used given the nature of the problem and also the prevailing contexts. Prioritization or rating of the different problem statements will be

3.1 **Brainstorming:** Brainstorming sessions will include researchers, stakeholders, and the SAFE target population. A session will involve verbally bouncing ideas off each other in the workshop in the hopes of finding a blended solution.
3.2 **Brain-writing:** Workshop participants will be requested to write down their ideas on the solution of SAFE implementation challenges before passing them on to someone else. For participants who are not able to write, the research assistants will encourage them to speak their ideas and they will help write them down and share with colleagues. The next person reads the ideas and adds their own, and so the process continues until each person’s ideas have done a full rotation. All ideas are then collected and placed in front of the group for discussion.
3.3 **Reverse thinking:** Reverse thinking is to flip the SAFE intervention problem on its head and come up with new ideas. Asking such question “how might we make our SAFE interventions more accessible?” could be changed to “how can we make it as difficult as possible for the SAFE target population to access SAFE interventions in this area?” The solutions that will come up from the reverse challenge would help the research team to envision what the opposite might be, leading closer to the solution needed to solve the SAFE intervention problems.

Ideation phase is followed by passing the ideas to the prototyping and testing phase and finalizing the tasks. If prototyping and testing phases prove that proposed novel SAFE interventions are implausible we will go back to the ideation phase.

## Phase 4: Prototype

At the prototyping phase, filtered problem statements and respective solution from ideation phase will be presented to the SAFE target population in succession to quickly test each design solution to address a defined SAFE implementation problem statement. Instantaneously, it will provide feedback to the research team on what the population think and feel about the new solutions including needed changes. The feedback will then be an important input to the research team in a workshop to revisit and further improve the intervention version or even decide to process another version which might be of higher preference and success among the SAFE target populations.

This phase will include conducting workshops that will involve the SAFE target population, researchers, and stakeholders who will be involved interactively to evaluate each version of the proposed innovation product until the acceptable version is reached.

**Objective:** Prototyping phase is to turn ideas from the ideation phase into something tangible which can be tested on real users. It is to identify solutions that have a high likelihood of solving SAFE implementation obstacles as being experienced by the SAFE target population.

The research team, stakeholders, and the SAFE target population will interact and assess each new version of the innovation which is provided as simple, scaled-down versions of the innovative solution to the prevailing SAFE implementation obstacles. The assessment will include recording observations, judging, and assessing the SAFE target population’s general behavior, interactions, and reactions to each version before making the final decision on the innovative solution to be submitted for testing. In addition, it will provide an opportunity for the research team to think critically about the anticipated innovative solutions and also an opportunity to fail quickly and cheaply so that less time and money is invested in a failing idea.

The prototype processing team will be composed of the SAFE target population, the research team, and co-opted stakeholders in a workshop setting where they will undertake participatory research that will jointly design and do the prototyping. This phase will enable researchers to get insights about innovative SAFE implementation intervention concerning its usability and SAFE target population experience early in the process.

the research team using accruing experiences will decide which of the innovative solutions will be suitable. This stage will assemble the ideas from all or some of the research notes, sketches, pictures, maps, and written versions of the point of view from the earlier phases to concretize it all and test it. Testing each proposed innovation will be based on the responses of the SAFE target population, stakeholders and research team to its suitability and acceptability and decide. In case of failure the process will appreciate that failing is part of learning, and failing is fine and should occur early but it needs to be analyzed to learn from the mistakes.

The workshop team envisages producing inexpensive, scaled-down versions of the innovative interventions generated in this phase, and demonstrates the functionality of the ideas before building the final SAFE intervention version. The SAFE target population will be actively involved in this component of designing the innovative intervention.

## Phase 5: Test phase

The design thinking process doesn’t follow a fixed sequence of steps, but the endpoint is testing phase which is to obtain a problem solution that is desirable, feasible, and viable.

**Objective:** To expose the human centered proposed novel intervention to wider group of the SAFE target population and observe how they interact with the product in its current state, and ask for feedback on how the experience and feels in order to confirm that it is feasible.

The rationale for conducting the testing on a prototype is to enable an understanding of how the SAFE target population will interact with the selected innovative solution to address the SAFE implementation problems in the local contexts. Our research team will conduct workshop(s), observations and one to one queries with the SAFE target population, leaders and other stakeholders to get their feed back to asses desirability, feasibility and viability of the proposed innovation which will be defined as follows:

a. Desirability - Proposed solution is desirable if it appeals to the needs, emotions, and behaviors of the SAFE target population.
b. Feasibility - Is the proposed solution technically possible?
c. Viability - is the solution self-sustaining?

However, when the process is completed a decision will be made to pack and adopt the SAFE intervention prototype for a rollout stage to a wider population at later times. However, in case the SAFE target population does not reveal desirability to the innovation, since design thinking is iterative and non-linear the process will resume the process with a different problem statement or solution ideas from the define or ideation phases. In that case, our research team will go back and re-start the define phase or empathy as the need arises until desirable and acceptable version is obtained.

## Data Collection Procedures

the study will employ a combination of qualitative data collection strategies including participant observations, photos/videos, IDI, KII and FGD among the SAFE target populations and related stakeholders. In addition, a series of workshops and community participatory approaches will be conducted at each phase to obtain the required data.

Two teams of 2 research assistants, and 1 ethnographer (one team for each study site /village) will be involved throughout the data collection process. In each study site, one overall supervisor will be identified to coordinate and oversee all daily data collection activities including handling logistics, providing feedback on a daily basis, managing collected data and providing support to the team in case of any challenges. The local coordinators from Trachoma control in each study site will the link the research team with the local leaders and SAFE target populations as well as to help with the translation during interviews.

Standard operating procedures (SOPs) for participant recruitment, data collection and data storage will be developed and implemented throughout the data collection period. All interviews and participants workshops will be audio recorded and notes taken. In addition, notes will be undertaken from participants’ observations and reflections from photos/videos as part of data collection.

## Capacity building of the research team and research assistants

Before commencement of the study, we will develop training manuals and provide training for research assistants, field supervisors, local coordinators, facilitators and the overall research team. The field team will include only those who demonstrate good performance during the training. During the training, the trainees will be provided with an overview of study and purposes, the design thinking approaches, ethnographic approaches, qualitative data collection and analysis strategies, professional conduct and research ethics, protection of those <18 years and other vulnerable populations, getting accustomed to data collections tools, field logistics, various SOPs for the project and covid-19 prevention protocol.

## Data Analysis and interpretation

Audio-recorded Kiswahili interviews and photovoice narratives will be transcribed to text in MS Word computer files and then translated into English. Additionally, all field notes from observations will be typed, and saved in MS Word computer files. We will conduct the initial transcript examination, to obtain the overall sense of the content of responses from participants regarding various issues. Analysis of data will follow qualitative thematic analysis[46] where the transcribed data in MS Word will be imported into NVivo 10 computer-assisted qualitative data analysis software. A codebook will be developed and shared among the research team. The team will discuss, refine and agree on a set of codes relevant to the objectives of the study. This discussion will provide an avenue for the addition of new codes, which may be incorporated into the codebook. The team will use negotiation to resolve disagreements about any issue concerning the identified codes and the coding process. Finally, a common codebook will be developed to facilitate data analysis. Each transcript will be coded to gather material into sub-themes, and finally core themes.

## Ethical clearance

This protocol will be submitted to the HKMU Institutional Ethics Review Board requesting a research ethical clearance. Thereafter the clearance will be submitted to the regional and district authorities in which the study sites are located. The authorities will also be requested to give the team permission to undertake the study as well as to introduce the team to the study sites, and health and governance officials. Each participant will be requested to give written consent and assent in a standard format as shown in Appendix A.

## Ethical considerations

The study will not involve any procedures to collect blood or any specimen for laboratory or other procedures hence no pain or injury will be inflicted on the study population. However, this study will use empathy methods that generate multi-dimensional data with the possibility of accessing or infringing on the privacy of individuals and or families. Data collectors will be trained and re-trained to make sure they observe and respect the privacy of the study subjects. The primary data will be kept strictly confidential and delinked from the data sources. Since the information will be used in each subsequent phase of the study, the data will be analyzed and synthesized before submission to the next phase in the process.

## Data Availability

It included literature reviews. the list is availale at the end of the documents

## Acknowledgement

We would like to express my profound and sincere gratitude to Bill and Melinda Foundation for accepting to fund this protocol

